# Outcome of Atrial Fibrillation in Obesity- Insights from The National Inpatient Sample Database

**DOI:** 10.1101/2025.02.12.25322152

**Authors:** Allan Santos Argueta, Archit Garg, Birgurman Singh, Olayiwola Olalekan Paul, Junaid Ali, Nirmal Kaur

## Abstract

**Background:** Obesity is considered a significant risk factor for numerous cardiovascular conditions. The prevalence of atrial fibrillation (AF) is elevated among patients with obesity. Weight loss has been shown to reverse cardiac remodelling, leading to lower recurrence of AF despite the better prognosis in obese patients.

**Methods:** We utilized the National Inpatient Sample 2016-2019 to extract patients ≥18 years of age admitted with AF as the primary diagnosis based on ICD 10 codes. We performed univariate and multivariate regression analysis for known coronary risk factors. We divided patients based on their body mass index (BMI), and our primary outcomes were determining the odds of electrical cardioversion (ECV) and cardiac ablation (CA) due to AF.

**Results:** The analysis included 1,625,809 weighted patients. Patients include underweight (6.66%), normal BMI (4.03), overweight (6.51%), obesity class I (20.65%), obesity class II (21.45%), and obesity class III (40.7).

After multivariate regression analysis, patients with obesity class I, II, or III had higher odds of ECV, irrespectively of coronary risk factors (OR 1.3, 95% CI 1.25-1.37, OR 1.3, 95% CI 1.32-1.43, OR 1.3, 95% CI1.29-1.38, respectively, with statistically significant P values).

However, underweight or normal BMI patients had fewer odds of ECV (OR 0.5 95%CI 0.49-0.61 and OR 0.6 95%CI 0.58-0.74, respectively, with P values <0.001). Meanwhile, there was no statistical significance between a BMI and the odds of CA.

**Conclusion:** Our study highlights the significant impact of BMI on managing AF, particularly regarding ECV. Patients in higher BMI categories (obesity class I to III) had increased odds of undergoing ECV, suggesting obesity influences treatment approaches and outcomes in AF management. Interestingly, BMI did not affect the likelihood of CA, indicating a complex relationship between body weight and AF treatment modalities warranting further investigation.

## Introduction

Obesity is a complex interplay of several cumulative biological, psychological, social, economic, and environmental factors. The pathways and mechanisms through which obesity leads to harmful health outcomes vary among individuals (1). WHO defines overweight and obesity as conditions characterized by the abnormal or excessive accumulation of body fat, which poses a significant risk to overall health (2). Obesity is assessed using the body mass index (BMI), which is calculated by using an individual’s weight (in kilograms) and height (in meters). A BMI of 30 or more is classified as obesity (3). It is estimated that between 39% and 49% of the global population (approximately 2.8 to 3.5 billion people) are affected by overweight or obesity. (4) Furthermore, findings from the Global Burden of Disease (GBD) study indicate a rising burden from elevated BMI; high BMI was noted to be responsible for approximately 4.0 million deaths in 2015. More than two-thirds of these deaths were attributed to cardiovascular diseases (CVD) (5). With increasing comorbidities, Atrial fibrillation (AF) is notably one of the most prevalent sustained arrhythmias. The growing burden of AF is attributed to many factors, such as an aging population, increasing prevalence of obesity, enhanced detection methods, and improved survival rates given medical advancement (6) (7).

Increased circulating blood volume due to obesity increases the workload of the left atrium (LA) and ventricle. This increased demand leads to chamber dilation and myocyte hypertrophy to sustain higher cardiac output. Over time, this leads to diastolic dysfunction, LA enlargement, and structural remodeling, all of which predispose to the risk of AF. A complex interplay of factors drives obesity-related AF, including hemodynamic and structural changes, electrical alterations, increased epicardial fat, myocardial fibrofatty infiltration, and metabolic dysfunction. In addition, neurohormonal dysregulation, oxidative stress, and altered ion channel expression also slow conduction velocity, alter action potentials and repolarization, reducing the effective refractory period, promoting ectopic activity, and hence AF (10). Multiple clinical studies have shown that weight reduction improves atrial fibrillation (AF) ablation outcomes by favorably altering structural and electrical remodeling. Bariatric surgery reduces the risk of new-onset AF and recurrence after ablation, alongside improvements in insulin resistance, blood pressure, and epicardial adipose tissue (EAT) volume. Observational studies and animal models have demonstrated that weight loss reduces LA EAT volume, myocardial fat infiltration, inflammation, and fibrosis. This reversal of obesity-induced remodeling reduces conduction abnormalities and improves atrial function (11) (12).

On the other hand, the “obesity paradox” suggests that overweight and obese patients with atrial fibrillation (AF) may have a lower risk of cardiovascular and all-cause mortality (13). However, the reasons for these controversial findings remain unclear, with randomized controlled trials supporting the paradox, while observational studies are more inconsistent. Despite this, evidence clearly shows that weight loss leads to a reduction in AF burden and reverses structural changes in overweight and obese patients. The 2024 ESC guidelines also recommend a 10% or more significant weight reduction to alleviate AF symptoms and burden (14). This study aims to evaluate the impact of BMI on the likelihood of undergoing electrical cardioversion and cardiac ablation in hospitalized atrial fibrillation patients.

## Methods

### Study Design and Data Source

We performed a retrospective cohort analysis using the Nationwide Inpatient Sample (NIS) database, focusing on data collected between 2016 and 2019. The National Inpatient Sample (NIS) is a publicly available, all-payer inpatient database in the United States, created under the Healthcare Cost and Utilization Project (HCUP) and funded by the Agency for Healthcare Research and Quality (AHRQ). It provides a comprehensive, nationally representative dataset of hospitalizations.

### Study Population and Variable Definitions

Adult patients (n=1,625,809) hospitalized with a primary diagnosis of atrial fibrillation (AF) were identified using the International Classification of Diseases, Tenth Revision, Clinical Modification (ICD-10-CM) code I48. We identified patients who underwent electrical cardioversion in this cohort using the procedure code 5A2204Z, while we identified those undergoing cardiac ablation using the procedure codes 02583ZZ and 02584ZZ.

Patients were further stratified by body mass index (BMI) categories, determined using ICD-10-CM codes for obesity and BMI classification. Additional variables extracted included age, sex, race, median household income quartile, hospital teaching status, bed size, location, and region. Comorbidities and overall severity of illness were assessed using the Charlson Comorbidity Index (CCI) score. {Table 1}

**Table 1:**
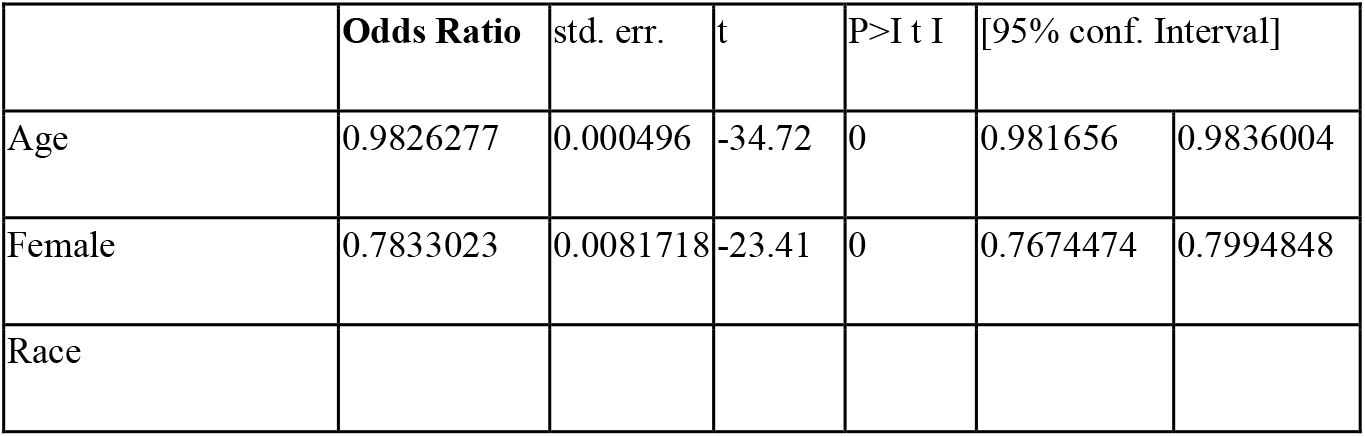
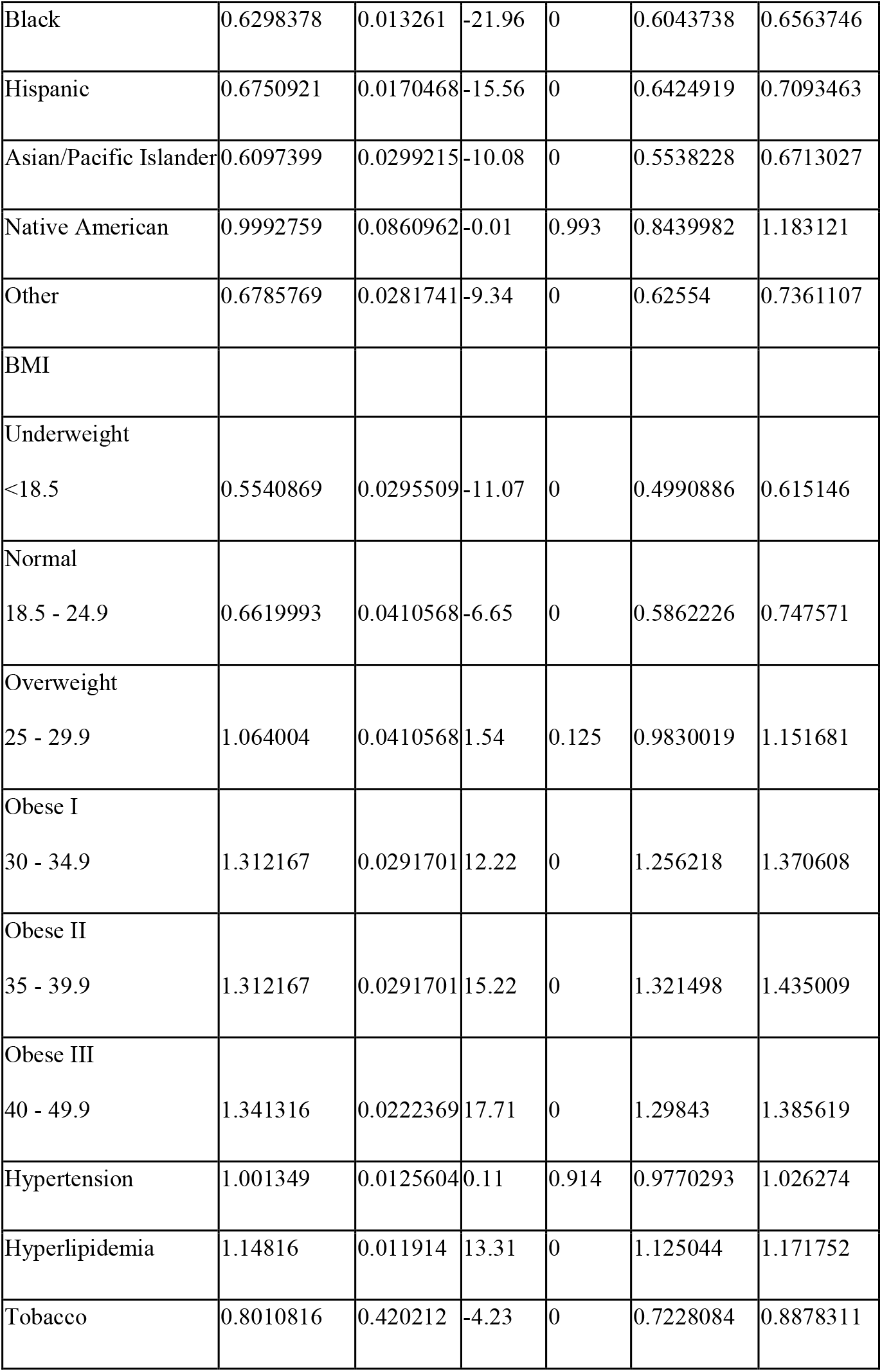
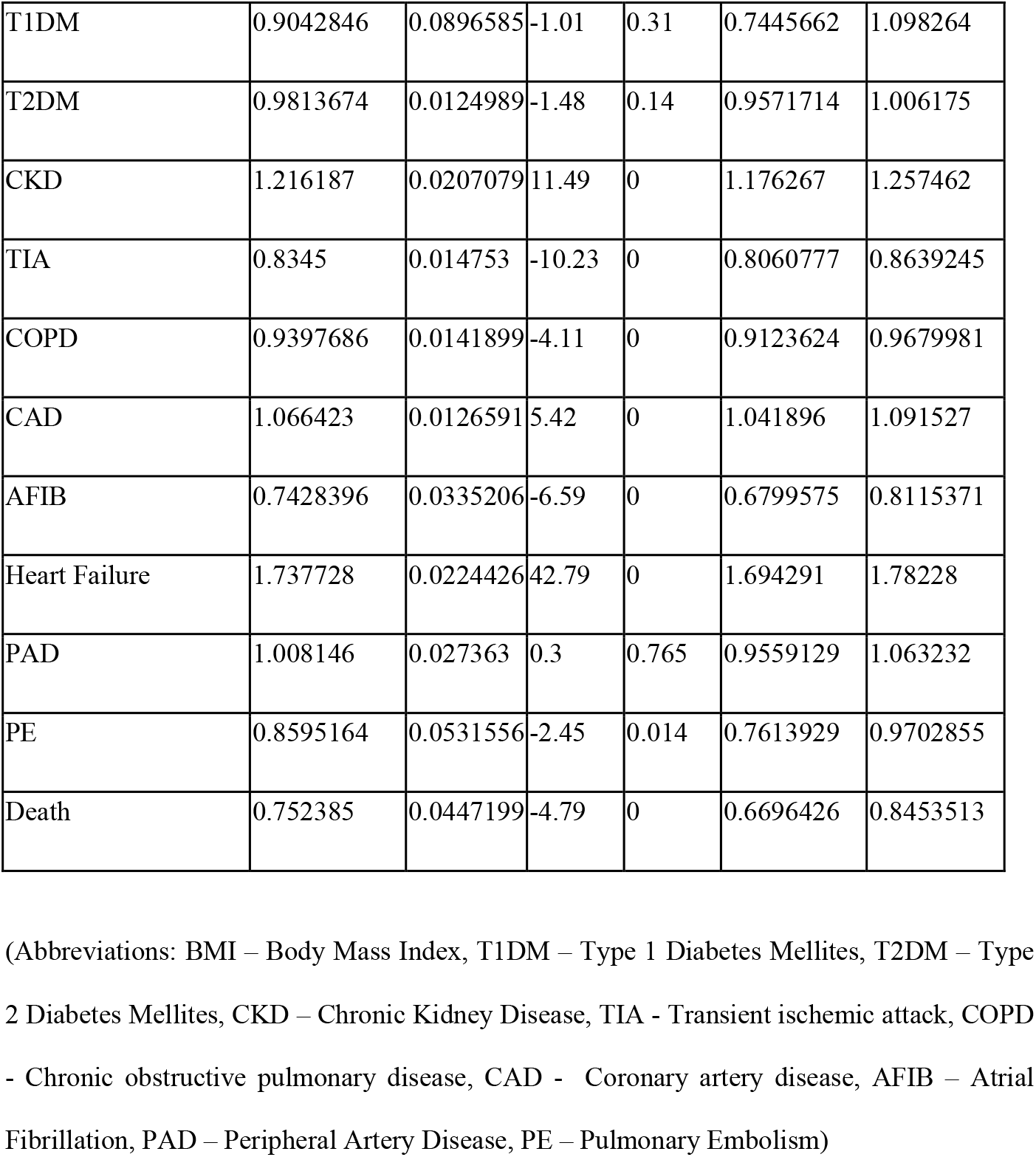
Patients Characteristics.

### Statistical Analysis

Univariate and multivariate regression analyses were performed to analyze the impact of BMI on the likelihood of undergoing electrical cardioversion and or cardiac ablation among hospitalized AF patients. In this study, we compared continuous variables using the Student’s t-test and categorical variables using the chi-squared test. We reported effect sizes as odds ratios (ORs) with 95% confidence intervals (CIs).

The study’s primary outcome was establishing an association between BMI and the odds of successful electrical cardioversion or cardiac ablation. Statistical significance was defined as a two-sided P-value <0.05. All analyses accounted for the NIS’s complex sampling design through appropriate weighting, stratification, and clustering using the Svy package in STATA version 17 (StataCorp LLC).

## Results

A total of 1,625,809 patients admitted to hospitals with atrial fibrillation (AF) from 2016 to 2019 were identified. The mean age was 70.53 years, with Caucasians making up 80.65% of the cohort. Most hospitalizations occurred in urban areas (89.29%), distributed regionally across the South (40.49%), Midwest (23.77%), Northeast (20.28%), and West (15.46%). Medicare was the most common insurance type (69.8%), followed by private insurers (20.6%). Patients were categorized by body mass index (BMI) as underweight (6.66%), normal BMI (4.03%), overweight (6.51%), obese class I (20.65%), obese class II (21.45%), and obese class III (40.7%).

In the cohort, 62,365 patients underwent cardiac ablation (3.84%), and 306,775 (18.87%) underwent electrical cardioversion. After adjusting for comorbidities, including hypertension, hyperlipidemia, diabetes mellitus, chronic kidney disease, cerebrovascular disease, chronic obstructive pulmonary disease, coronary artery disease, heart failure, peripheral artery disease, and pulmonary embolism, a higher BMI was significantly associated with increased odds of electrical cardioversion. Patients classified as obese class I, II, and III had adjusted odds ratios (OR) of 1.3 (95% CI 1.25–1.37), 1.3 (95% CI 1.32–1.43), and 1.3 (95% CI 1.29– 1.38), respectively (P < 0.001 for all). In contrast, underweight and normal BMI patients had lower odds of electrical cardioversion (OR 0.5, 95% CI 0.49–0.61 and OR 0.6, 95% CI 0.58– 0.74, respectively; P < 0.001 for both).

BMI was not significantly associated with outcomes across all categories in patients who underwent cardiac ablation. Underweight patients (OR 1.01, 95% CI 0.85-1.2, p=0.85), those with normal BMI (OR 0.81, 95% CI 0.64-1.02, p=0.08), and overweight patients (OR 1.05, 95% CI 0.89-1.23, p=0.52) showed no statistically significant differences. Similarly, obesity demonstrated no significant association, including Class I (OR 0.99, 95% CI 0.91-1.09, p=0.96), Class II (OR 1.02, 95% CI 0.93-1.12, p=0.56), and Class III (OR 0.95, 95% CI 0.88-1.02, p=0.17).

No statistically significant correlation was observed between BMI and the likelihood of undergoing cardiac ablation. {Figure 1}

**Figure 1:**
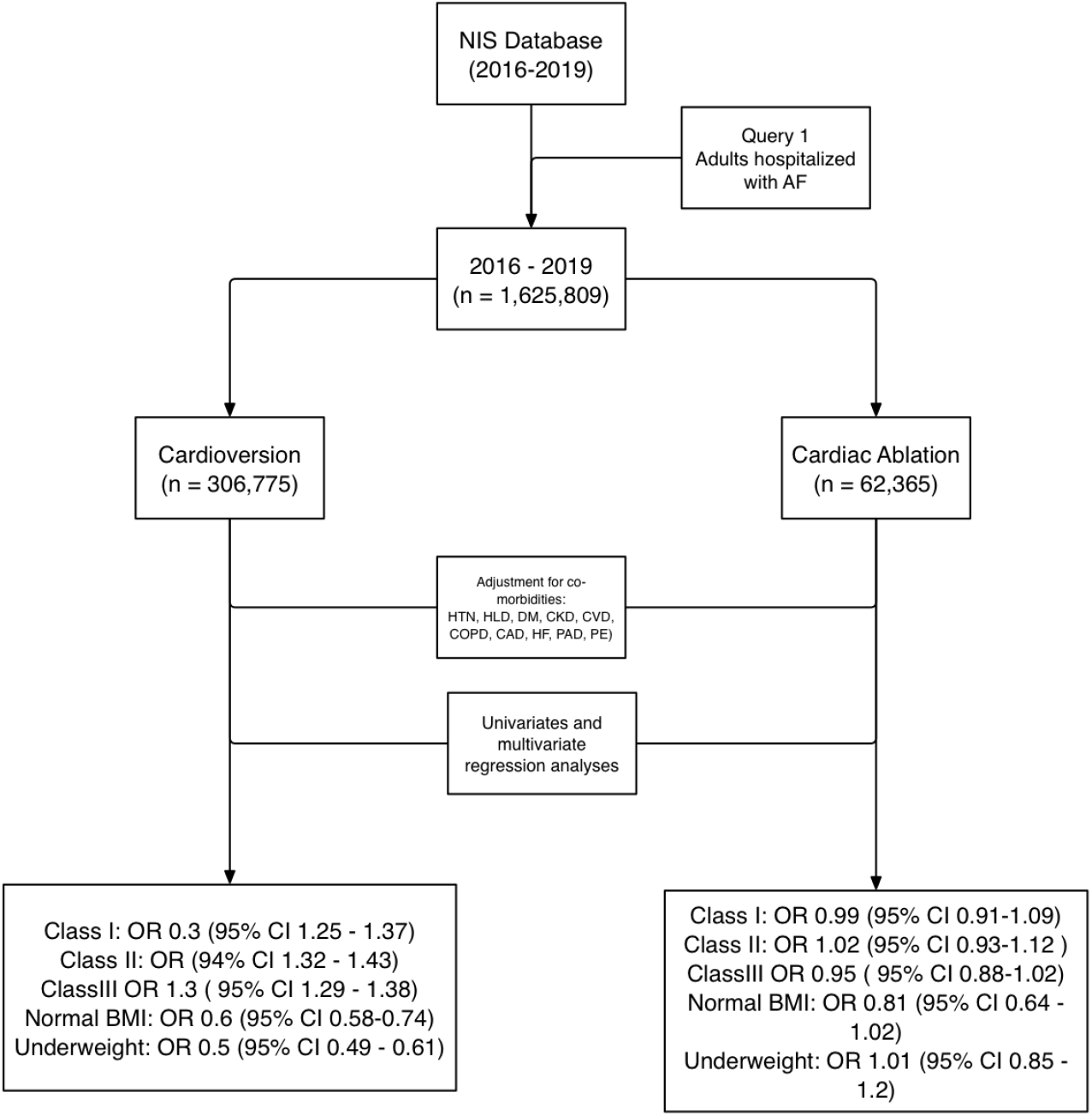
Flowchart

## Discussion

The incidence and prevalence of obesity and atrial fibrillation (AF) have risen in parallel, both exhibiting exponential growth over time. According to the World Obesity Atlas 2023, it is estimated that by 2030, 78% of the US adult population will be overweight/obese. [15] Similarly, it is estimated that by 2030, AF is expected to increase by 23%, with an annual growth rate of 4.6%. [16] By 2050, the prevalence of AF will be 16 million in the US and 16-17 million in Europe. [16] Obesity is increasingly recognized as a risk factor for AF, with the incidence rates of AF being 50% higher among the obese population. [17] Hence, obesity, often referred to as a “global pandemic,” has sparked growing concern about the critical role of weight reduction in protecting heart health.

Our study showed that obesity classes I, II, and III had statistically increased odds of undergoing electrical cardioversion in comparison to people with normal BMI or those who are underweight. The odds ratio for electrical cardioversion amongst people with obesity class I, II, and III was 1.3 (95% CI 1.25 – 1.37), 1.3 (95% CI 1.32 – 1.43), and 1.3 (95% CI 1.29 – 1.38), respectively (P < 0.001 for all). In contrast, the odds ratio for electrical cardioversion among subjects with normal BMI and underweight was 0.5 (95% CI 0.49 – 0.61) and 0.6 (95% CI 0.58 – 0.74), respectively; P < 0.001 for both. Our study also found no statistically significant relationship between obesity and the likelihood of undergoing cardiac ablation. This might suggest that BMI might impact the management of AF in hospital settings. Our study is unique in that it not only highlights that increased BMI is associated with increased occurrence of AF but also signifies the impact of BMI on the choice of management for AF. To the best of our knowledge, this is the first NIS study assessing the outcomes of AF in obese patients stratified on the basis of BMI and how it influences the management of AF.

Our study demonstrates that among the patients undergoing electrical cardioversion, the odds of AF were significantly higher among patients who were obese. The result was statistically significant among all the obesity classes (30% increased association among all classes). In contrast, the odds of AF were significantly lower among patients with normal BMI or underweight (50% lower in normal BMi and 40% lower in underweight patients). Prior studies have also reported similar findings. Wong et al., in their meta-analysis involving 51 studies, noted that for every 5 kg/m2 increase in BMI, the incidence of AF increases by 10-29% increased risk of incident, post-operative, and post-ablation AF. [18] Similarly, the HUNT3 study reported 18% and 59% increased AF risk among overweight and obese subjects, respectively. [19] A recently published meta-analysis by Folli et al. involving 50 studies reported an increased risk of newly diagnosed AF among overweight, obese, and morbidly obese patients and recurrent post-ablation AF among obese and morbidly obese patients. [13] This suggests that an increase in BMI is directly proportional to the risk of both new onset and recurrent AF.

The data regarding AF incidence and BMI is conflicting, with some studies reporting increased AF incidence with obesity and others supporting the hypothesis called the “obesity paradox”. As per the paradox, higher BMI is inversely associated with mortality rates in AF. [20] Rodriguez-Reyes et al. reported that high BMI and waist-hip ratio were associated with lower case fatality rates among AF patients. [21] Similarly, in the Gulf SAFE trial, AF patients with higher BMI had lower risks of stroke, bleeding, heart failure admission, and all-cause mortality. [22] A likely explanation for the lower all-cause mortality rates could be more aggressive treatment and management of comorbidities for obese patients. For example, the ARISTOTLE trial showed that the use of statins and beta blockers was 50% and 68% for obese AF patients compared to 34% and 56% for non-obese AF patients. [10] Although these studies report the rate of complications, especially all cause of mortality is lesser during follow up periods, the rates of prevalence and post-ablation recurrence of AF still has a direct proportional relationship with BMI. [10, 22, 23] Hence, our article strengthens the weight reduction point to decrease the AF rates.

Ligero et al., in their study, reported that increased BMI (≥25 kg/m2) was independently associated with an increased risk of recurrence of AF after initial intervention with electrical cardioversion. [24] This shows that BMI plays a role in AF recurrence. Interestingly, in our study, no significant association was observed between AF and obesity classes among patients undergoing cardiac ablation. This might indicate that BMI affects the treatment of AF. Patel et al., in their study, reported increased failure rates of catheter ablation among obese patients. [25] Similarly, Chilukuri et al., in their prospective study, reported an increase in BMI by one unit and increased the recurrence rates of AF by 11% after catheter ablation. Moreover, catheter ablation can be technically more challenging in obese patients. Hence, electrical cardioversion might be tried first in patients with higher BMIs.

Our study has several limitations. First, the NIS database’s cross-sectional and administrative design limited the collection of patient-level information essential for classifying patient severity, such as laboratory, radiographic, and echocardiographic results. Furthermore, the identification of AF types, such as new onset or paroxysmal, was made more difficult by the errors in ICD-10 coding. In addition, the indication of choosing cardiac ablation and electrical cardioversion was not specified. Moreover, this study does not assess the long-term outcomes of AF and cannot comment on the recurrence rates of AF and whether or not it is associated with BMI. Furthermore, the database’s emphasis on in-hospital events raises the possibility of overlooking post-hospitalization outcomes, such as out-of-hospital sudden cardiac death, long-term mortality, and complications. Despite these limitations, our study has the advantage of being an extensive retrospective analysis with a good sample size and reporting AF association stratified based on BMI and classified based on cardiac intervention undertaken for the AF.

In conclusion, our study highlights the significant impact of BMI on managing atrial fibrillation, particularly among patients undergoing electrical cardioversion (ECV). Obesity may affect treatment strategies and results in the management of atrial fibrillation (AF), as patients in higher BMI categories (obesity classes I, II, and III) showed higher odds of undergoing ECV. Interestingly, BMI did not affect the probability of cardiac ablation (CA), suggesting a more nuanced relationship between body weight and AF treatment options that needs more research. These results highlight the significance of tailored treatment plans with an emphasis on weight control to maximize results.

## Data Availability

All data produced in the present study are available upon reasonable request to the authors

## Acknowledgments

None

